# Toric Intraocular Lens Implantation in Cataract Patients with Corneal Opacity

**DOI:** 10.1101/19003319

**Authors:** Ho Ra, Hyun Seung Kim, Man Soo Kim, Eun Chul Kim

## Abstract

**Aims:** To evaluate the effect of toric intraocular lens implantation in cataract patient with corneal opacity and high astigmatism.

**Methods:** 31 eyes of 31 patients who underwent cataract surgery with toric intraocular lens implantation were included. All patients had corneal opacity with regular astigmatism. Preoperative total corneal astigmatism was determined considering posterior astigmatism using a rotating Scheimpflug camera (Pentacam®: Oculus, Wetzlar, Germany). At 2 months after toric intraocular lens implantation, we evaluated residual astigmatism, uncorrected visual acuity (UCVA) and best corrected visual acuity (BCVA).

**Results:** Postoperative UCVA and BCVA (0.30 ± 0.17, 0.22 ± 0.16LogMAR) statistically improved compared to preoperative UCVA and BCVA (1.2 ± 0.34, 1.1 ± 0.30LogMAR, respectively) (P<0.01). Postoperative residual refractive astigmatism (1.2 ± 0.35D) was statistically reduced compared to preoperative refractive astigmatism (2.4 ± 0.65D) (P<0.05). Preoperative and postoperative total corneal astigmatism values were not statistically different. All cases achieved visual acuity were as good as or better than that preoperatively. The percentage of corneal opacity covering pupillary area had significant negative correlation with postoperative UCVA and BCVA (logMAR) (R=-0.88 P<0.00001 and R=-0.87 P<0.00001, respectively)

**Conclusion:** Toric intraocular lens implantation can improve UCVA, BCVA, and refractive astigmatism in cataract patient with corneal opacity. The percentage of central corneal opacity covering pupillary area is the major prognostic factor for postoperative visual improvement. Therefore, toric intraocular lens implantation should be considered for cataract patients who have corneal opacity with high astigmatism.

## Introduction

Corneal opacity and cataract are the primary causes of decreasing visual acuity. There are two currently surgical treatments options for patients with corneal opacity and cataract. The Triple procedure, simultaneous penetrating keratoplasty (PK), cataract extraction and intraocular lens (IOL) implantation provides a shorter visual rehabilitation period.^1^ However, the disadvantages include risk of expulsive hemorrhage, inadequate cortical cleaning and inaccuracy in IOL power calculation can decrease postoperative visual acuity.^2^ Cataract surgery without PK is sometimes associated with good visual acuity when the corneal opacity partially obscures the pupillary area.^3^ Even though visual outcomes after cataract surgery in eyes with corneal opacities can vary, corneal opacity severity may be one of the major prognostic factors of visual acuity.^4^ When there is opacity in the patient’s cornea, astigmatism usually occurs in the vertical axis (with-the-rule astigmatism), horizontal axis (against-the-rule astigmatism), oblique axis, or irregular axis. When the patient’s cornea has regular astigmatism, good visual acuity can be achieved through astigmatic correction. During cataract surgery, astigmatism can be corrected by prescription glasses, contact lenses, corneal relaxing incisions, astigmatic keratotomy, limbal relaxing incisions, excimer laser ablation, or toric IOL implantation.^5^ Toric IOL implantation is the most reliable and effective method for correcting regular astigmatism during cataract surgery. We hypothesized that toric IOL implantation can improve visual acuity in patients with cataract and regular corneal astigmatism.

To the best of our knowledge, there are currently no studies that have evaluated the efficacy of toric IOL implantation in cataract patients with corneal opacity.

## Materials and methods

This study was conducted by performing a retrospective chart review and data analysis. This study was conducted in compliance with Institutional Review Board regulations, informed consent regulations, sponsor and investigator obligations, and the Declaration of Helsinki. The Institutional Review Board (IRB) / Ethics Committee of Bucheon St. Mary’s Hospital approved this study protocol.

### Patients

Thirty-one patients that had cataract and corneal opacity with regular astigmatism were enrolled from Bucheon St. Mary’s Hospital from June 2017 to April 2018.

Inclusion criteria were corneal opacification that involved the visual axis and advanced cataract in patients with regular corneal astigmatism and eyes with partially visible anterior capsules and pupillary margins. Patients with a history of any ocular injury or disorder, infection, inflammation, surgery within the prior 6 months and an eye with irregular astigmatism were excluded.

### Preoperative evaluation

All patients underwent a complete preoperative ophthalmological examination. The demographic and perioperative data were recorded. Uncorrected and corrected distance visual acuity were expressed as logMAR. Manifest refraction, biometry and keratometry with the IOLMaster partial coherence interferometry device (Carl Zeiss Meditec AG), corneal topography to exclude irregular astigmatism, slit lamp examination, and dilated funduscopy were performed. The IOL manufacturer’s web-based toric calculator was used to determine the required cylinder power and axis for implantation. The total corneal astigmatism was measured using the Scheimpflug system (Pentacam^®^, Oculus, Germany). The percentage of corneal opacity covering pupillary area was calculated using image J (National Institutes of Health, Maryland).

### Operative Procedures

Before surgery, the corneal limbus was marked at the 0°, 90°, and 180° meridians with the patient in a sitting position after instilling topical anesthetic eye drops. All operations were performed under topical anesthesia by a single skilled surgeon (E.C.K) using the Intrepid Infiniti system (Alcon Laboratories, Inc., Fort Worth, TX, USA). The corneal steep axis and 6.0 mm ring were marked with gentian violet. Surgery was performed through a clear corneal incision at the steep astigmatic axis. After topical ocular anesthesia was applied, a 2.75 mm clear corneal incision was made using a 2.75 mm double-blade keratome (Alcon). Sodium hyaluronate 1.0% (Hyal Plus, LG Life Science, Seoul, Korea) was used to reform and stabilize the anterior chamber. A continuous curvilinear capsulotomy was made with a 6.0 mm corneal marker using Inamura capsulorhexis forceps (Duckworth & Kent Ltd., Baldock, UK). Hydrodissection and hydrodelineation were achieved using a balanced salt solution. Phacoemulsification was performed using 2.75-mm-sized phaco-tips and infusion/aspiration (I/A) cannulas in the micro- and small-incision groups, respectively. A clear preloaded IOL (Tecnis ZCT; Abbott Medical Optics) was implanted in the capsular bag. The wound was not sutured (Video). Postoperative treatment consisted of gatifloxacin 0.3% (Gatiflo, Handok, Chungbuk, Korea) and fluorometholone acetate 0.01% (Ocumetholone, Samil, Seoul, Korea) eye drops four times a day for four weeks.

### Statistical Analysis

Statistical analysis was performed using a commercial program (SPSS for Windows; version 21.0.1; SPSS Inc., Chicago, IL). The Wilcoxon signed rank test was used to compare pre- and postoperative BCVA and refractive and keratometry astigmatism. P values < 0.05 were considered statistically significant.

## Results

Thirty-one eyes of 31 patients were enrolled in the study. Table 1 summarizes the patient demographics and the preoperative data.

**Table 1.**
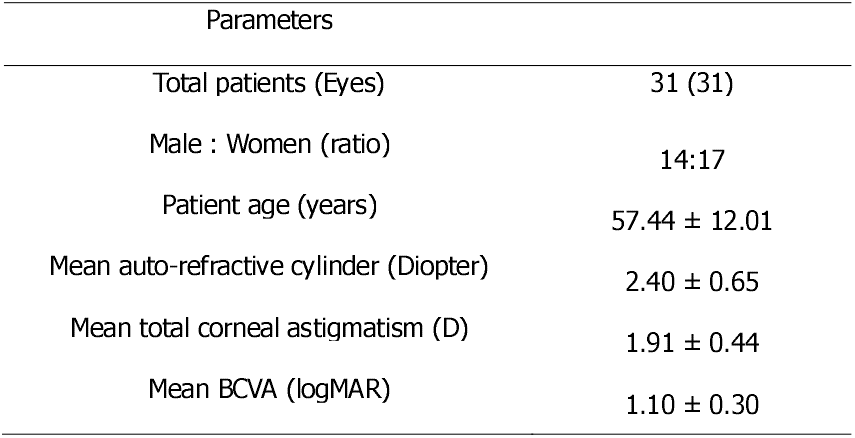
The patient demographics and the preoperative data.

| Parameters |  |
| --- | --- |
| Total patients (Eyes) | 31 (31) |
| Male : Women (ratio) | 14:17 |
| Patient age (years) | 57.44 ± 12.01 |
| Mean auto-refractive cylinder (Diopter) | 2.40 ± 0.65 |
| Mean total corneal astigmatism (D) | 1.91 ± 0.44 |
| Mean BCVA (logMAR) | 1.10 ± 0.30 |

### Data represent mean± standard deviation

The preoperative diagnoses comprised trichiasis, corneal ulcer, herpes keratitis, previous pterygium surgery, uveitis, traumatic corneal scar, and unknown (Table 2). Postoperative UCVA (0.30 ± 0.17) and BCVA (0.22 ± 0.16) were significantly improved compared to preoperative UCVA (1.20 ± 0.34) and BCVA (1.10 ± 0.30) (P<0.05) (Figure 1). The UCVA 2 months postoperatively was 20/32 or better in 19 eyes (61.3%) and 20/25 or better in 7 eyes (22.6%) (Figure 2). The postoperative residual refractive astigmatism (1.20 ± 0.35 D) was statistically reduced compared to preoperative refractive astigmatism (2.4 ± 0.65 D) (P<0.05). Preoperative and postoperative total corneal astigmatism (1.91 ± 0.44 & 1.52 ± 0.38 D, respectively) were not statistically different (Figure 3). The percentage of residual astigmatism within ± 0.5D was 22.6% (7 eyes of 31), and that within ± 1.0D was 58.1% (27 eyes of 31) (Figure 4).

**Table 2.** The causes of corneal opacity

|  | Number of eyes |
| --- | --- |
| Trichiasis | 3 |
| Corneal ulcer | 5 |
| Herpes keratitis | 2 |
| Previous pterygium surgery | 3 |
| Uveitis | 5 |
| Traumatic corneal scar | 6 |
| Unknown | 7 |
| Total | 31 |

**Figure 1.**
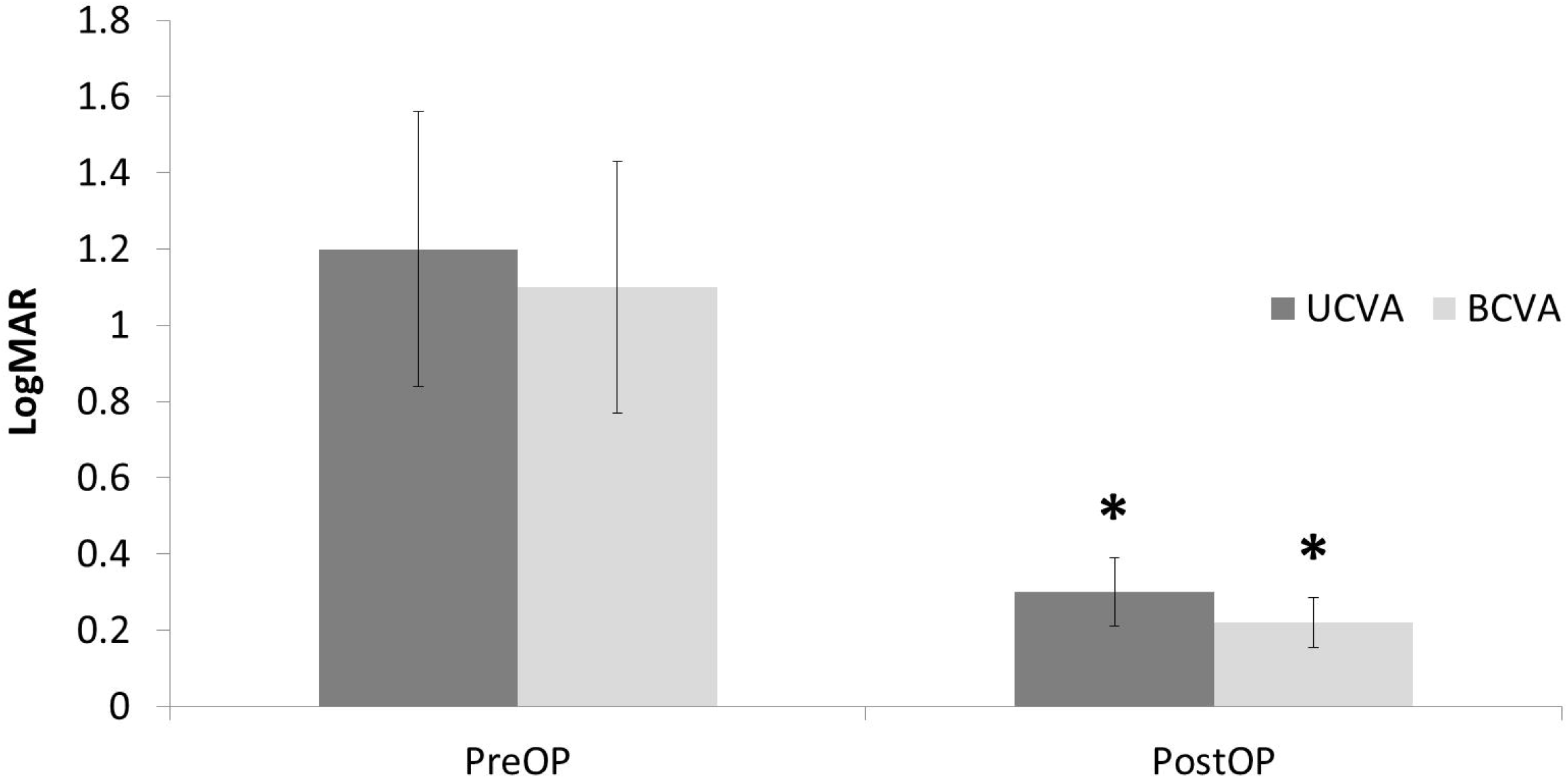
Preoperative and 2 month postoperative visual acuity. Values are presented as mean±SD. UCVA: uncorrected visual acuity, BCVA: best corrected visual acuity Postoperative UCVA (0.30 ± 0.17) and BCVA (0.22 ± 0.16) were significantly improved compared to preoperative UCVA (1.20 ± 0.34) and BCVA (1.10 ± 0.30) (P<0.05).

**Figure 2.**
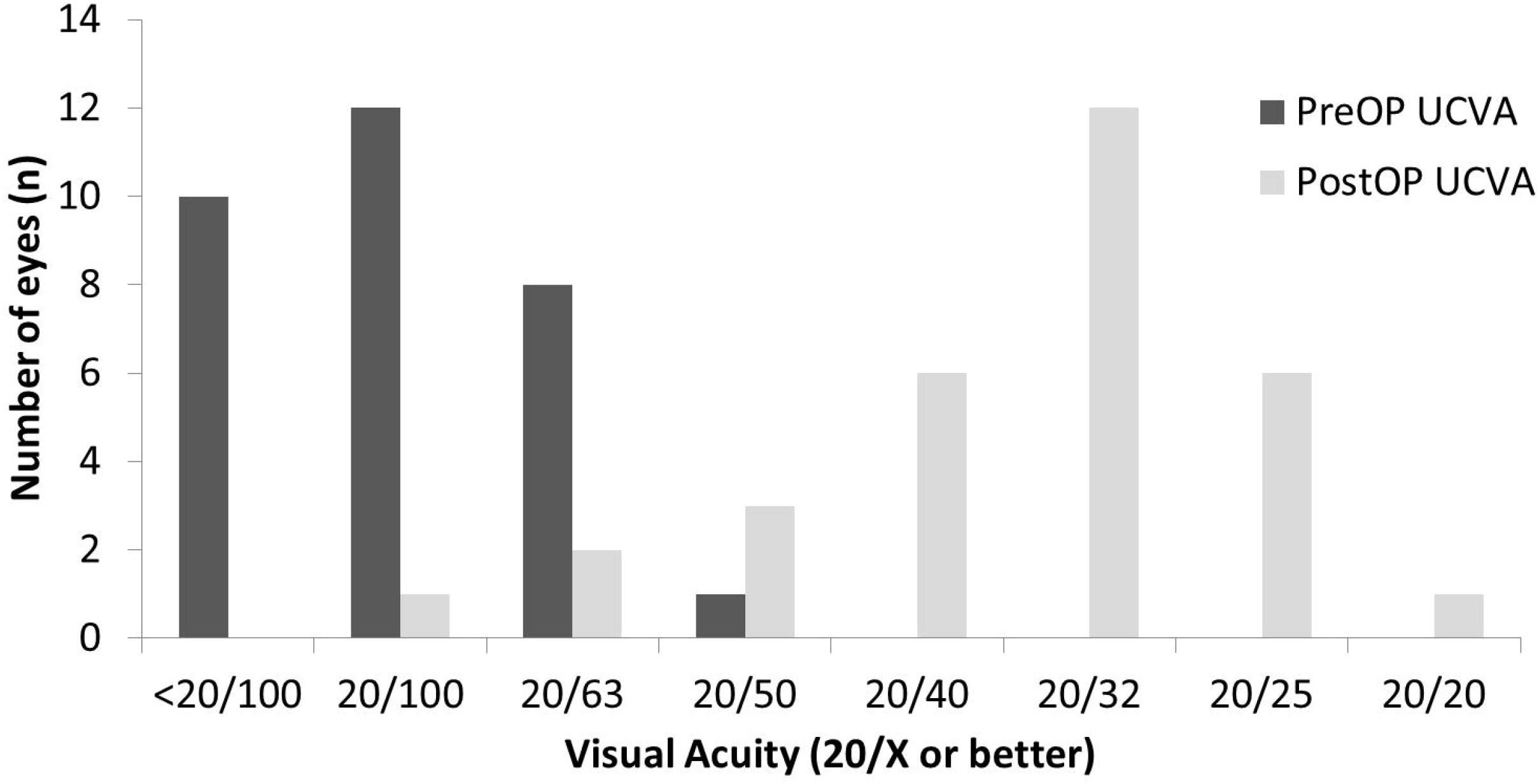
Preoperative and 2 month postoperative visual acuity distribution. Values are presented as mean±SD. UCVA: uncorrected visual acuity The UCVA 2 months postoperatively was 20/32 or better in 19 eyes (61.3%) and 20/25 or better in 7 eyes.

**Figure 3.**
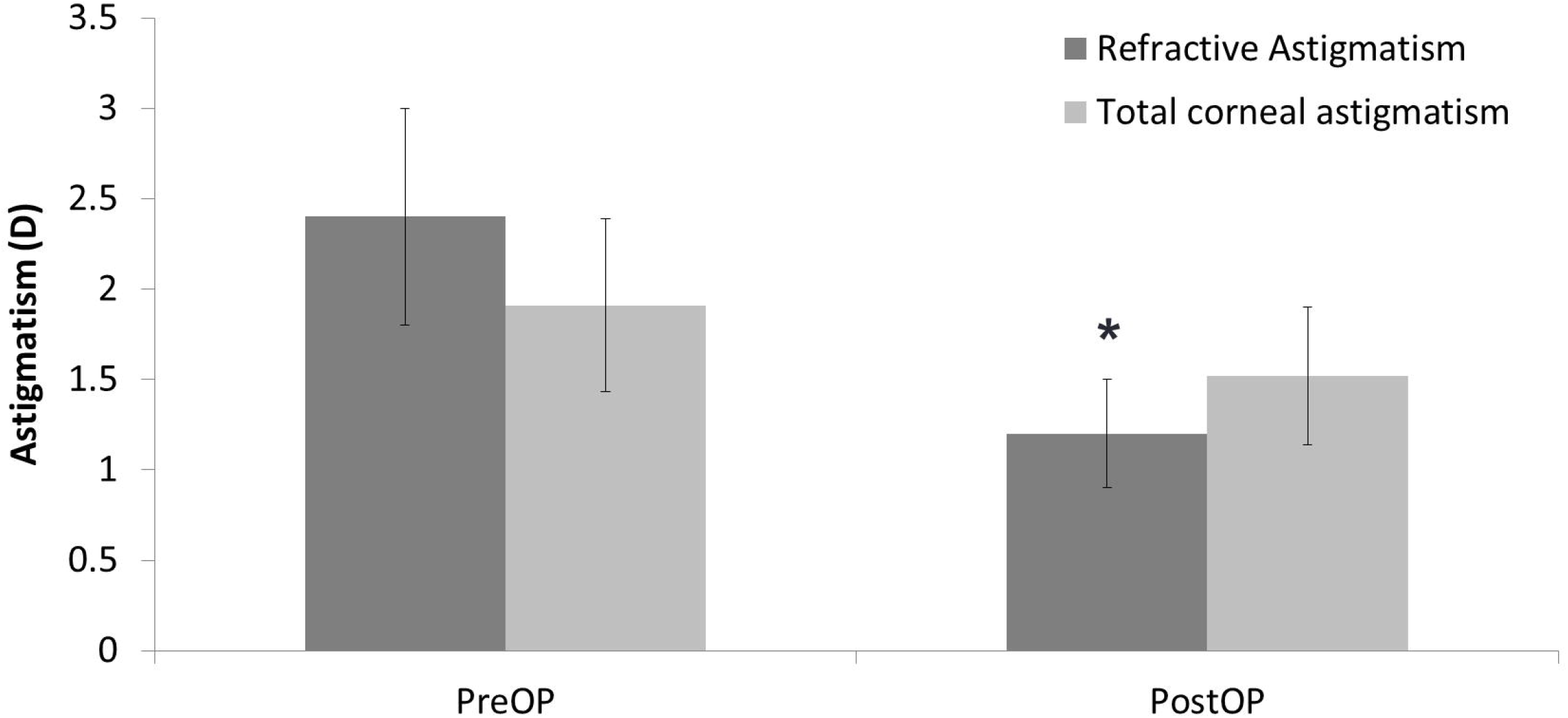
Preoperative and 2 month postoperative astigmatism. Values are presented as mean±SD. D: Diopter Postoperative residual refractive astigmatism (1.20 ± 0.35 D) was statistically reduced compared to preoperative refractive astigmatism (2.4 ± 0.65 D) (P<0.05).

**Figure 4.**
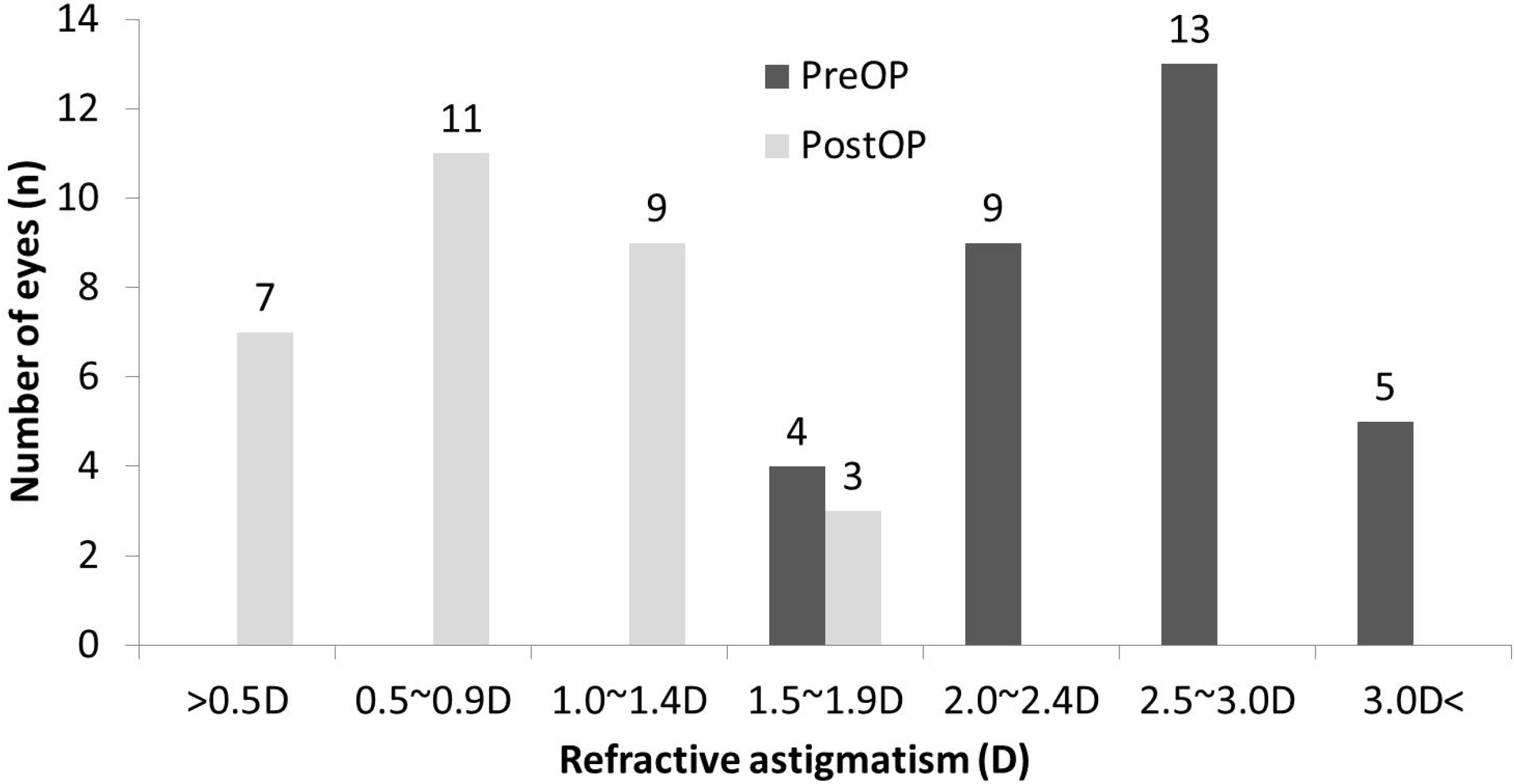
Preoperative and 2 month postoperative astigmatism distribution. Values are presented as mean±SD. D: Diopter The percentage of residual astigmatism within ± 0.5D was 22.6% (7 eyes of 31) and within ± 1.0D was 58.1% (27 eyes of 31).

**Figure 5.**
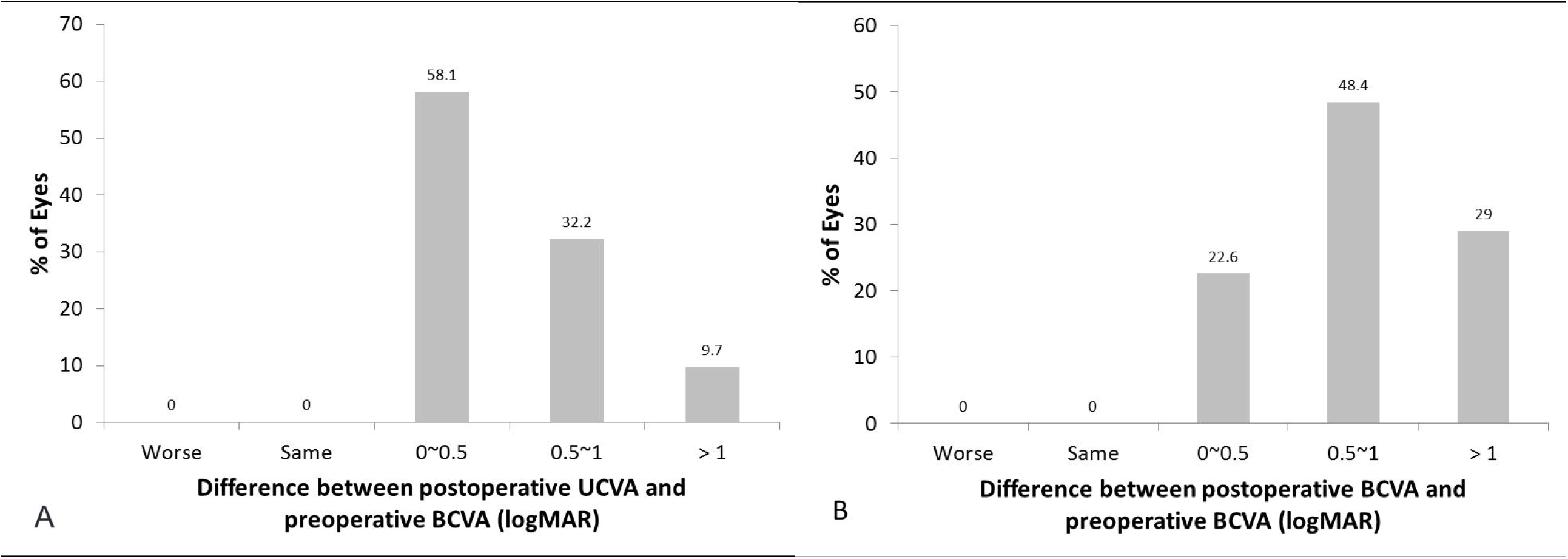
Comparison of preoperative BCVA and postoperative UCVA(A) & BCVA(B) in logMAR. **Values are presented as mean±SD**. **UCVA: Uncorrected visual acuity** **BCVA: Best corrected visual acuity** 100% cases achieved UCVA and BCVA were as good as or better than that preoperatively with correction.

100% cases achieved UCVA and BCVA were as good as or better than that preoperatively with correction. 42.9% cases achieved better UCVA compared to preoperative BCVA 0.5 logMAR over and 77.4% cases achieved better BCVA compared to preoperative BCVA 0.5 logMAR over (Figure 4 A & B). The percentage of corneal opacity covering pupillary area had significant negative correlation with postoperative UCVA and BCVA (logMAR) (R=-0.88 P<0.00001 and R=-0.87 P<0.00001, respectively)

Figure 7 shows data from a 77-year-old male patient treated for bacterial keratitis in his right eye. His inferior temporal cornea was opaque and very thin (Figure 7B) because of previous bacterial keratitis (Figure 7A). His preoperative BCVA was 20/200 and refractive astigmatism was 2.5D. After a Tecnis ZCT225 was inserted, postoperative 2 month UCVA was 20/22, BCVA was 20/20, and residual refractive astigmatism was 1.0D. Figure 8 involves a 63-year-old female patient who reported central corneal opacity in her right eye from birth. Her preoperative BCVA was 20/60 and refractive astigmatism was 2.75D. After a Tecnis ZCT300 was inserted, her postoperative 2 month UCVA was 20/24, BCVA was 20/22, and residual refractive astigmatism was 0D.

**Figure 6.**
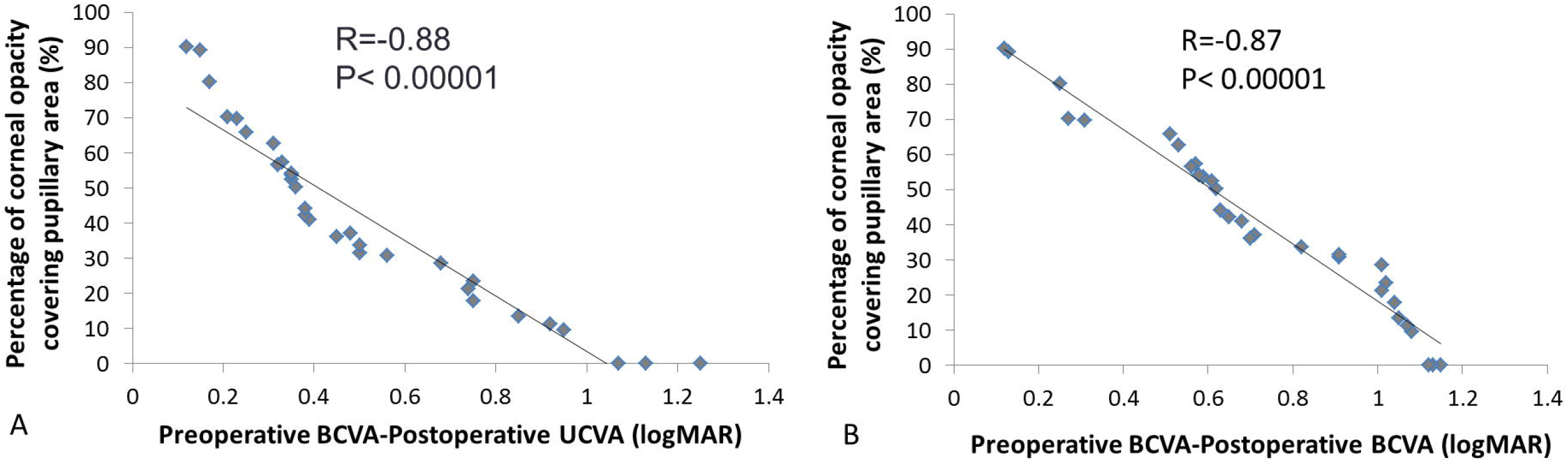
Correlation of the percentage of corneal opacity covering pupillary area and postoperative UCVA(A) and BCVA(B) (logMAR). **UCVA: Uncorrected visual acuity** **BCVA: Best corrected visual acuity** The percentage of corneal opacity covering pupillary area had significant positive correlation with postoperative UCVA and BCVA (logMAR) (R=0.88 P<0.00001 and R=0.87 P<0.00001, respectively)

**Figure 7.**
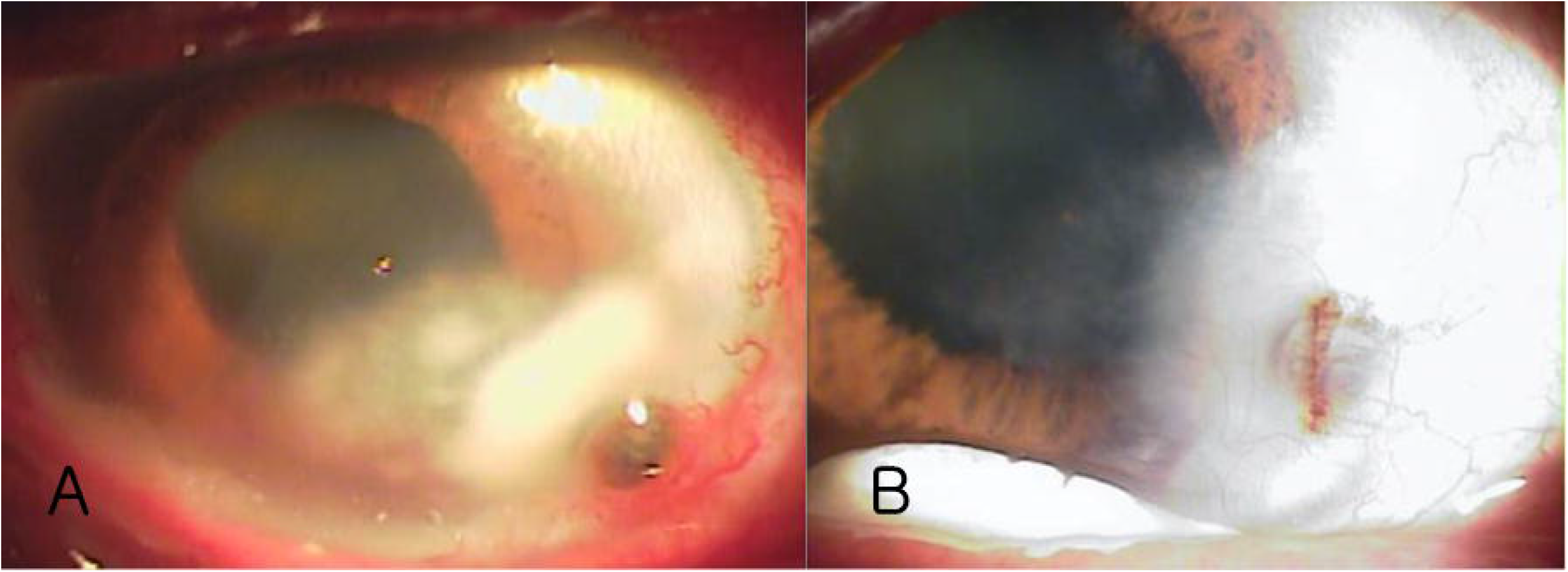
Preoperative image of a patient treated for bacterial keratitis. Inferior temporal cornea is opaque and very thin (Figure 5B) because of previous bacterial keratitis (Figure 5A). The patient’s preoperative UCVA, BCVA, and refractive astigmatism improved at 2 months postoperatively.

**Figure 8.**
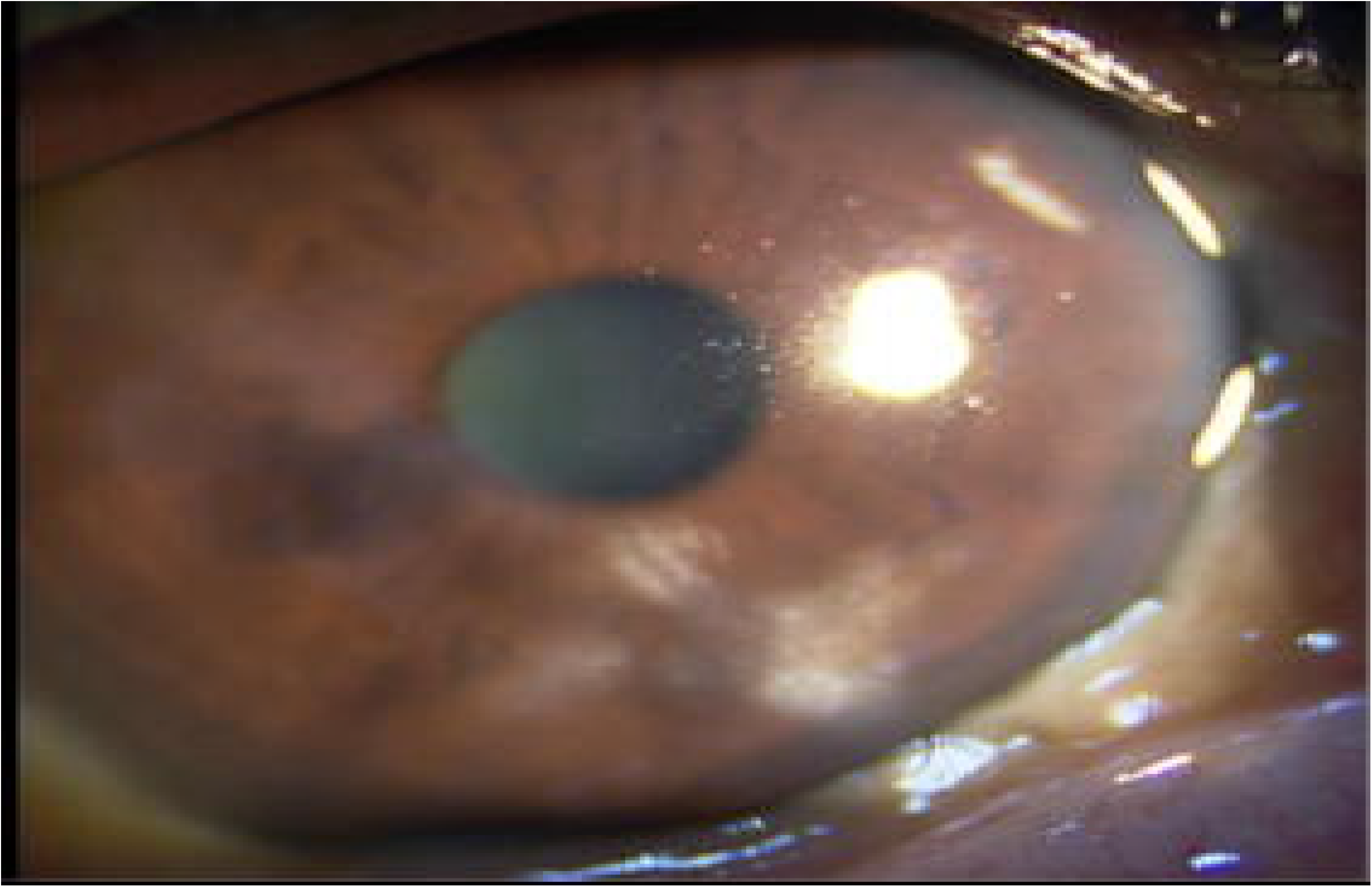
Preoperative image of a patient with congenital central corneal opacity. The patient’s preoperative UCVA, BCVA, and refractive astigmatism were improved at 2 months postoperatively.

## Discussion

About 60% of patients with normal corneas that undergo cataract surgery have more than 0.75D of corneal astigmatism.^6^ Therefore, patients with cataract and corneal opacity have greater corneal astigmatism than the normal population.^7^ Consequently, simultaneous penetrating keratoplasty and cataract surgery may be the preferred treatment option in this patient group.^2 8^ However, simultaneous penetrating keratoplasty and cataract surgery may induce more astigmatism than the preoperative condition.^2 8^ Phacoemulsification and IOL implantation in selected cases of coexisting cataract and corneal opacity are safe and can provide suboptimal but long-term vision when penetrating keratoplasty is not an option or there is a high risk of graft failure.^4^ Cataract surgery with toric IOL implantation could better improve visual acuity than simultaneous penetrating keratoplasty and cataract surgery when the central cornea is not totally opaque.^3^ BCVA improvements after surgery are less likely for more severe opacity indices that involve the pupil, according to the reflectivity signal.^4^

In this study, postoperative UCVA (0.30 ± 0.17) and BCVA (0.22 ± 0.16) were significantly improved compared to preoperative UCVA (1.20 ± 0.34) and BCVA (1.10 ± 0.30) (P<0.05). Ho et al. reported preoperative mean UCVA and BCVA of 20/800 and 20/630, respectively, which significantly improved to 20/200 and 20/160 (P < 0.001) after cataract surgery with monofocal IOL implantation in patients with corneal opacity.^4^ In the Ho et al. study, visual acuity improvement was less than that in our study, because astigmatism was not corrected in this study.

Müftüoğlu İK reported that mean preoperative BCVA significantly increased (0.7 ± 0.3 [range: 0.3–1.3] logMAR to 0.1 ± 0.04 [range: 0.05–0.15] logMAR; P<0.05) at a mean of 8.71 ± 64.11 months after cataract surgery with toric IOL implantation in patients with cataract formation and high astigmatism after penetrating keratoplasty.^9^ Development of cataract is highly possible after PK because of chronic high-dose steroid use and surgical intervention. Toric IOLs are reported to be an effective modality to correct astigmatism in patients with cataract.^10-12^ Additionally, visual acuity improvement was greater in the study by Müftüoğlu İK than in our study, because the corneas in their study were relatively clear after PK compared to those in our study..

In this study, the 2 month postoperative UCVA was 20/32 or better in 19 eyes (61.3%) and 20/25 or better in 7 eyes (22.6%). This indicates that the majority of patients who underwent toric IOL implantation will not need to wear glasses daily. The corrected distance Snellen visual acuity (with spectacles or contact lenses) 12 months postoperatively was 20/32 or better in 82% of eyes in keratoconus patients with toric IOL implantation.^13^ In the normal cataract patients that did not have corneal opacity, mean LogMAR UDVA and BDVA were 0.19 ± 0.12 and 0.14 ± 0.10, respectively. In addition, a postoperative UDVA of 20/40 or better was achieved in 92.6% of eyes.^14^

In this study, postoperative residual refractive astigmatism (1.20 ± 0.35 D) was statistically reduced compared to preoperative refractive astigmatism (2.4 ± 0.65 D) (P<0.05). Müftüoğlu İK reported that the mean preoperative corneal keratometric astigmatism was 5.4 ± 0.9 D (range: 4.25–7.00 D) at the corneal plane and 6.3 ± 1.0 D (range: 4.9–8.1 D) at the spectacle plane, and the average manifest refractive astigmatism was 1.5 ± 0.7 D (range: 0.25–2.25 D) at postoperative month 1 after toric IOL implantation in patients that had previously undergone penetrating keratoplasty.^9^ Postoperative refractive astigmatism significantly decreased in their study, which was in agreement with the results of our study. In contrast, refractive astigmatism persisted after cataract surgery with monofocal IOL implantation.^4^ In normal cataract patients without corneal opacity, mean refractive cylinder decreased significantly from −3.73 ± 1.96 to −1.42 ± 0.88D (*p*< 0.001), while keratometric cylinder did not change significantly (*p*= 0.44) after toric IOL implantation.^14^ The visual and refractive astigmatic outcomes were comparable to reports on normal cataract patients without corneal opacity, even though there was corneal opacity in this study.

In other study, 92.3% cases achieved visual acuity were as good as or better than that preoperatively with correction after monofocal IOL implantation.^4^ However, in this study, all cases achieved visual acuity were as good as or better than that preoperatively with correction after toric IOL implantation.

In central corneal opacity, phacoemulsification should be performed when the extent of opacity is small enough to improve visual acuity after surgery.^15^ In this study, we included patients with peripheral corneal opacity as demonstrated in Figure 7, and large central corneal opacity as in Figure 8. Patient with peripheral corneal opacity had more improved postoperative UCVA and BCVA compared to patient with central corneal opacity, in these cases. Regarding visual outcome, Ho Y. et al. reported that there is no significant correlation between logMAR BCVA and corneal densitometry and OCT grading (*p*>0.05).^4^ However, in this study, the percentage of corneal opacity covering pupillary area was negatively correlated with postoperative UCVA and BCVA improvement (P<0.05) (Figure 6).

Therefore, we concluded that not the central corneal opacity size, but the percentage of central corneal opacity covering pupillary area was the major prognostic factor for postoperative visual improvement. Patients with paracentral corneal opacity are the best candidates for cataract surgery to optimize vision.^15^ But, if corneal opacity did not cover the whole central pupillary area, good vision can be achieved with cataract surgery and toric IOL implantation.

This was the first study to evaluate toric IOL implantation outcomes in cataract patients with corneal opacity. The postoperative visual acuity was significantly improved in spite of the previously existing corneal opacity. The postoperative residual refractive astigmatism was also significantly improved. Postoperative UCVA and BCVA were correlated with the percentage of corneal opacity covering pupillary area. Therefore, toric IOL implantation is effective for improving visual acuity in patients with corneal opacity and cataract.

### Study Limitations

The short duration of follow up (2 month) and lack of control over the monofocal IOL of the subjects were among the limitations of this study. A multicenter clinical trial with a larger sample size and longer follow up period is suggested to observe the long-term efficacy of the toric intraocular lens implantation in cataract patients with corneal opacity.

## Data Availability

All data relevant to the study are included in the article or uploaded as supplementary information

## Data Availability

All data relevant to the study are included in the article or uploaded as supplementary information

## Video legend

**Video. Implantation of Toric Intraocular Lens in Patient with Cataract and Corneal Opacity**.

Toric Intraocular Lens was implanted in patient with central corneal opacity.

## Reference

1 Javadi MA, Feizi S, Moein HR. Simultaneous penetrating keratoplasty and cataract surgery. J Ophthalmic Vis Res 2013;8:39–46.

2 Shimmura S, Ohashi Y, Shiroma H, et al. Corneal opacity and cataract: triple procedure versus secondary approach. Cornea 2003;22:234–8.

3 Panda A, Krishna SN, Dada T. Outcome of phacoemulsification in eyes with cataract and cornea opacity partially obscuring the pupillary area. Nepal J Ophthalmol 2012;4:217–23.

4 Ho YJ, Sun CC, Chen HC. Cataract surgery in patients with corneal opacities. BMC Ophthalmol 2018;18:106.

5 Agresta B, Knorz MC, Donatti C, et al. Visual acuity improvements after implantation of toric intraocular lenses in cataract patients with astigmatism: a systematic review. BMC Ophthalmol 2012;12:41.

6 Hoffmann PC, Hutz WW. Analysis of biometry and prevalence data for corneal astigmatism in 23,239 eyes. J Cataract Refract Surg 2010;36:1479–85.

7 Bouzas A, Marcakis G. [Correlation between corneal leukoma and induced astigmatism]. Bull Mem Soc Fr Ophtalmol 1986;97:201–3.

8 Hayashi K, Hayashi H. Simultaneous versus sequential penetrating keratoplasty and cataract surgery. Cornea 2006;25:1020–5.

9 Muftuoglu IK, Akova YA, Egrilmez S, et al. The Results of Toric Intraocular Lens Implantation in Patients With Cataract and High Astigmatism After Penetrating Keratoplasty. Eye Contact Lens 2016;42:e8–e11.

10 Mendicute J, Irigoyen C, Aramberri J, et al. Foldable toric intraocular lens for astigmatism correction in cataract patients. J Cataract Refract Surg 2008;34:601–7.

11 Chang DF. Comparative rotational stability of single-piece open-loop acrylic and plate-haptic silicone toric intraocular lenses. J Cataract Refract Surg 2008;34:1842–7.

12 Bauer NJ, de Vries NE, Webers CA, et al. Astigmatism management in cataract surgery with the AcrySof toric intraocular lens. J Cataract Refract Surg 2008;34:1483–8.

13 Mol IE, Van Dooren BT. Toric intraocular lenses for correction of astigmatism in keratoconus and after corneal surgery. Clin Ophthalmol 2016;10:1153–9.

14 Lubinski W, Kazmierczak B, Gronkowska-Serafin J, et al. Clinical Outcomes after Uncomplicated Cataract Surgery with Implantation of the Tecnis Toric Intraocular Lens. J Ophthalmol 2016;2016:3257217.

15 Sharma N, Singhal D, Maharana PK, et al. Phacoemulsification with coexisting corneal opacities. J Cataract Refract Surg 2019;45:94–100.

